# Beta regression with spatio-temporal effects as a tool for hospital impact analysis of initial phase epidemics: the case of COVID-19 in Spain

**DOI:** 10.1101/2020.06.27.20141614

**Authors:** Gil Jannes, Jesús Barreal

## Abstract

COVID-19 has put an extraordinary strain on medical staff around the world, but also on hospital facilities and the global capacity of national healthcare systems. In this paper, Beta regression is introduced as a tool to analyze the rate of hospitalization and the proportion of Intensive Care Unit admissions over both hospitalized and diagnosed patients, with the aim of explaining as well as predicting, and thus allowing to better anticipate, the impact on hospital resources during an early-phase epidemic. This is applied to the initial phase COVID-19 pandemic in Spain and its different regions from 20-Feb to 08-Apr of 2020. Spatial and temporal factors are included in the Beta distribution through a precision factor. The model reveals the importance of the lagged data of hospital occupation, as well as the rate of recovered patients. Excellent agreement is found for next-day predictions, while even for multiple-day predictions (up to 12 days), robust results are obtained in most cases in spite of the limited reliability and consistency of the data.

## 1 Introduction

COVID-19, the disease caused by the SARS-CoV-2 virus, has led to one of the largest pandemics known in recent times. It has caused an effective breakdown of many sectors of the economy and of most social activities, as well as an evidently tremendous concern for the health sector, from hospitals over pharmaceutical companies to scientific institutions. From a hospitalary point of view, three types of cases can be distinguished: asymptomatic and mild cases which do not require hospital treatment, hospitalized patients or inpatients, and patients that require admission to an Intensive Care Unit (ICU).

The large number of patients in the third group is especially problematic for hospitals, because they lead to a sharp peak in the demand for hospital beds, equipment and medical attention, and thus to a saturation of the ICUs’ available space and resources, both material and human. The current crisis has shown that the national and global healthcare and economic systems were insufficiently prepared to supply the demand for medical equipment and staff required to cope with this situation. These issues are ultimately related to the number of infected, and in particular those that will require hospitalization (regular or ICU). Many studies therefore focus on analysing absolute numbers and patterns of spread, see e.g. Dong et al. (2020), Liu et al. (2020), Bi et al. (2020); while health management studies in the context of COVID-19 focus mainly on qualitative aspects, such as the design of an emergency plan [Cao et al. (2020)], adaptation of Hospital facilities and the importance of communication with staff [Grange et al. (2020)], or on analysing the risk for hospital personnel [Cheung (2020) and Yu et al. (2020)] or for other hospital visitors, such as pregnant women [Chen et al. (2020)].

Here, a complementary point of view is presented, allowing for a quantitative analysis and prediction of the *ratio* or proportion of patients that require hospital and/or ICU admission. The main advantage of these ratios as compared to absolute numbers is that they are scale-independent indicators of the strain put on the installations, amount of beds and other resources of hospitals of any size in regions with any amount of population. We mention two key applications here. First, this method allows to forecast which proportion of hospital resources and capacity should be reserved for ICU purposes during such crises across all hospitals of a particular region. The scale-independence of the ratios means that it can be applied equally to hospitals of all sizes. Second, several countries have made use of temporary hospitals created exclusively for COVID-19 patients. The ratios that we will study, and in particular the UCI/Hosp ratio, provide a very simple and powerful way of planning the distribution of such temporary facilities.

The purpose of this paper is thus to provide policymakers in the health sector with statistical and econometric tools to predict hospital occupation rates, both general and at the ICU level, during the first phase of future pandemics with features similar to the current COVID-19 pandemic. These predictions will relie on the identification of temporal patterns and spatial effects in the ratios of hospitalized patients over total cases, ICU over hospitalized, and ICU over total cases. At the heart of the methodology lies the well-known Beta regression, which is ideally suited for modeling rates and proportions, and adds several elements of information to the more common predictions based on logistic distributions, such as Villalobos-Arias (2020), Buizza (2020), Bliznashki (2020), Wu et al. (2020) and Huang et al. (2020), or the Weibull distribution, as in Zhang (2016). Beta distributions have been applied before to the health sector, in Gange et al. (1996), Hunger et al. (2012) and Moraes et al. (2012), but for quite different purposes.

A three-step methodology will be developed. First, a Beta distribution will describe the probability function for all possible values of the ratios of interest; second, a logistic distribution will be used to estimate the time evolution of raw data of diagnosed, hospitalized, ICU patients, and recovered patients; third and foremost, a Beta regression will be used to develop a prediction method for the ratios described earlier based on their lagged values and on the proportion of recovered patients.

To illustrate this methodology, it will be applied to Spain during the early-phase COVID-19 epidemic. There are several reasons why Spain is an interesting example. First, Spain has been one of the hardest hit countries so far, and thus provides a sample of a statistically interesting size. Second, parts of the Spanish hospital system have indeed been overwhelmed, which is precisely the focus of this research. Third, Spain’s regional division in autonomous communities, each responsible for the collection and communication of the COVID-19 data, means that Spain naturally provides a relatively wide and diverse geographical sample. In this sense, spatial patterns will be essential to describe the evolution of the hospital ratios in the Spanish health system. The paper therefore develops distribution models for both Spain globally (in the main text) and for its regions separately (in the Appendices). The methodology is robust and could be applied to other geographical areas.

The paper is structured as follows. In the next section, a data description is given to visualize the early-phase Spanish pandemic evolution, both temporal and spatial. The following section describes the statistical methods and the econometric models, with an emphasis on the Beta regression with spatial and temporal components. Some key results with regard to the current COVID-19 data for Spain are discussed next. Additional information about the methods and the data, as well as detailed results for the Spanish regions separately, are given in a series of Appendices.

## 2 Data and variables

The Spanish Ministry of Health records and provides daily data about the pandemic evolution in each of the Spanish regions [Ministerio de Sanidad (2020)]. Figure 1 shows the mean number of newly recorded cases, hospitalized patients and fatalities per 100 000 inhabitants per day in each of the Spanish regions from 20-Feb-2020 to 08-Apr-2020, counted from the first non-zero day. The plots clearly show the inhomogeneous character of the spatial distribution for all three variables. Madrid (MD), Catalonia (CT), the Basque Country (BC) and La Rioja (RI) are the areas with the highest impact in proportion to their number of inhabitants, while the South, the North-West and the Spanish Islands present lower values. Note that for the first group (“cases”) we take the data initially provided by the Spanish Ministry of Health at face value, regardless of whether these cases have been effectively clinically diagnosed through a medical test, or are just based on a symptomatic analysis. This remark is important because the first category (positive medical test) could include asymptomatic patients, whereas the second category (symptomatic analysis) could include false negatives, for example due to influenza or other diseases. In fact, the uncertainty about the true number of people affected by COVID-19 is one of the major current incognita. Coma et al. (2020) suggest that several thousand COVID-19 cases in Catalonia alone may have been misdiagnosed as influenza. For Italy, Modi et al. (2020) estimate that the true number of COVID-19 might be at least twice the official number, while Vollmer & Bommer (2020) estimate a factor of possibly 10 to 15 worldwide. An advanced mathematical model for undetected infections and their influence on the spread of the COVID-19 virus has been developed in Ivorra (2020).

**Figure 1:**
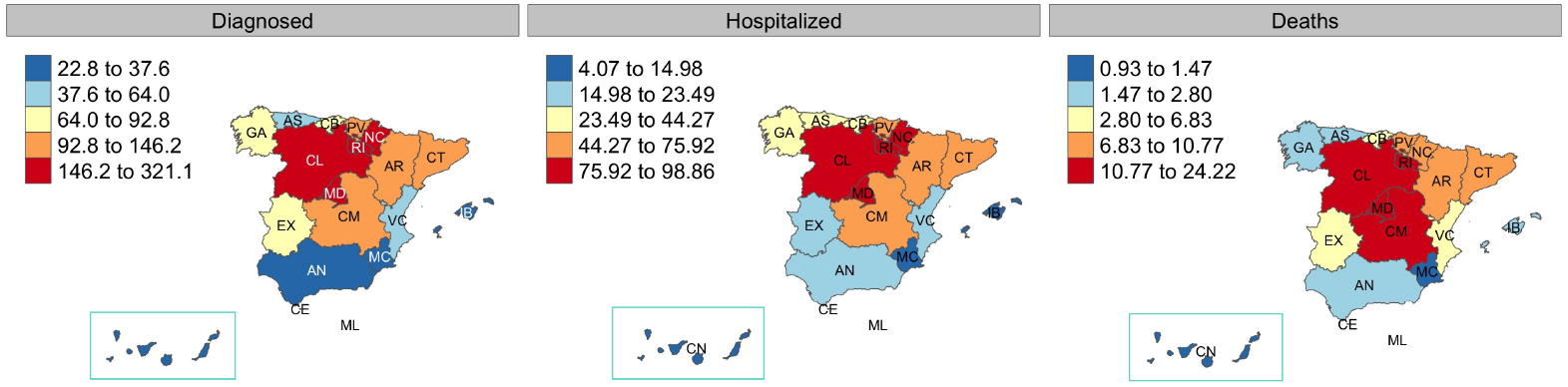
Absolute number of cases, hospitalized and deaths related with COVID-19

Figure 2 shows the temporal evolution of cases, inpatients, ICU patients, deaths, and recovered patients in absolute numbers for each Spanish region, plotted from the day of the first official case recorded (which can differ by various days from region to region). While the number of cases follows a qualitatively similar evolution in all regions, there are some interesting differences in the evolution of the number of hospitalized patients. In particular, in CM (Castilla-la-Mancha) and Madrid (MD) these had already started to go down by April 8th, while the other regions were still in an increasing phase.

**Figure 2:**
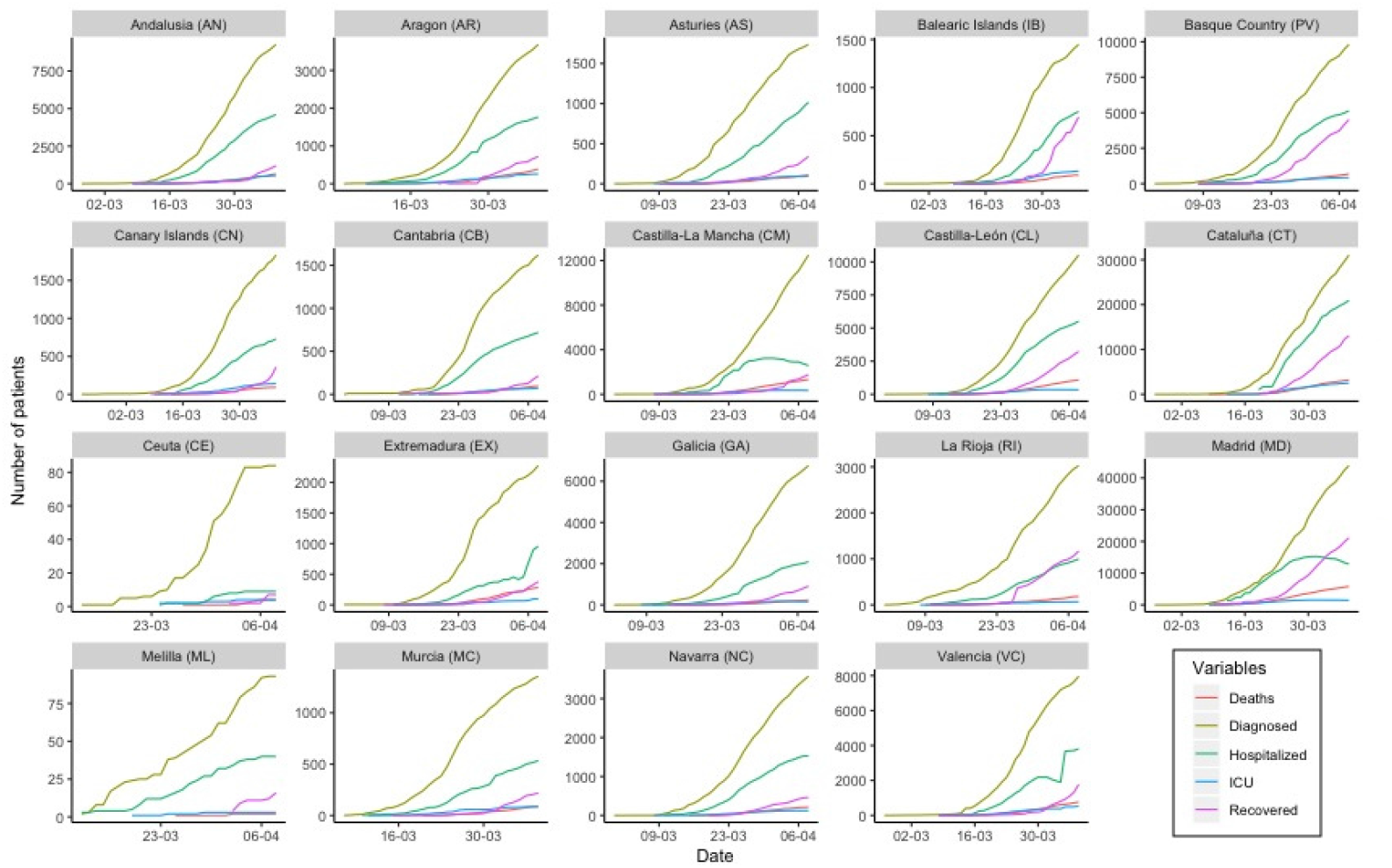
Absolute number of diagnosed cases, hospitalized, ICU patients, recovered patients and fatalities related with COVID-19 in the different Spanish regions.

As stated in the Introduction, this paper aims to provide policymakers and hospital managers with some statistical methods to manage future similar situations, depending on the strain put on ICUs. We will from now on focus on the proportion of hospitalized patients to total cases (1a), the proportion of ICU over total hospitalized patients (1b), and finally the proportion of ICU patients over total cases (1c), since these are the variables crucial for the research goal. At the global (Spanish) level, these variables are defined as

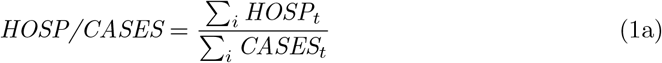

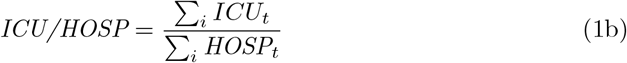

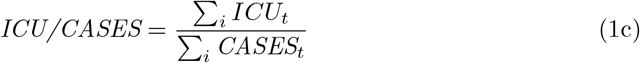

where the summation is over the regions, and the index *t* indicates that these are calculated day by day. Note that the third variable can in principle be obtained as the product of the previous two.

As mentioned before, there is a large uncertainty about the real number of infected, so the description “cases” should be taken with caution, since this number can fluctuate a lot depending on whether one refers to “infected” (but not necessarily ill), “symptomatically diagnosed” or “clinically tested”, and in the latter case: through PCR or antibody testing. Current data on the number of cases is therefore rather unreliable and inconsistent, since different countries or regions use different methodologies. In any case, the methodology that we will present is independent of how the cases are counted. As we will show, it allows an accurate prediction of the three proportion variables just defined.

Figure 3 shows the geographical distribution of the mean of these three variables over the period considered (starting from the first case recorded until April 8th). The first map shows the ratio of hospitalized over total cases, and shows that regions such as Madrid (MD), Castilla-León (CL), Catalonia (CT) and the Basque Country (PV) present a higher rate of hospitalization than Extremadura (EX), Galicia (GA) or La Rioja (RI). The second map shows the severity of the hospitalized cases, i.e. the ratio of ICU patients over total hospitalized patients, with high recordings in Murcia (MC) and the Canary (CN) and Balearic (BI) Islands, and low values in center-north: Castilla-León (CL), La Rioja (RI) and the Basque Country (PV). The last plot involves ICU cases over total cases, which is crucial to estimate hospital strains. Catalonia (CT), Aragon (AR) and the Canary (CN) and Balearic (BI) Islands record the highest values; Galicia (GA), Extremadura (EX) and La Rioja (RI) the lowest.

**Figure 3:**
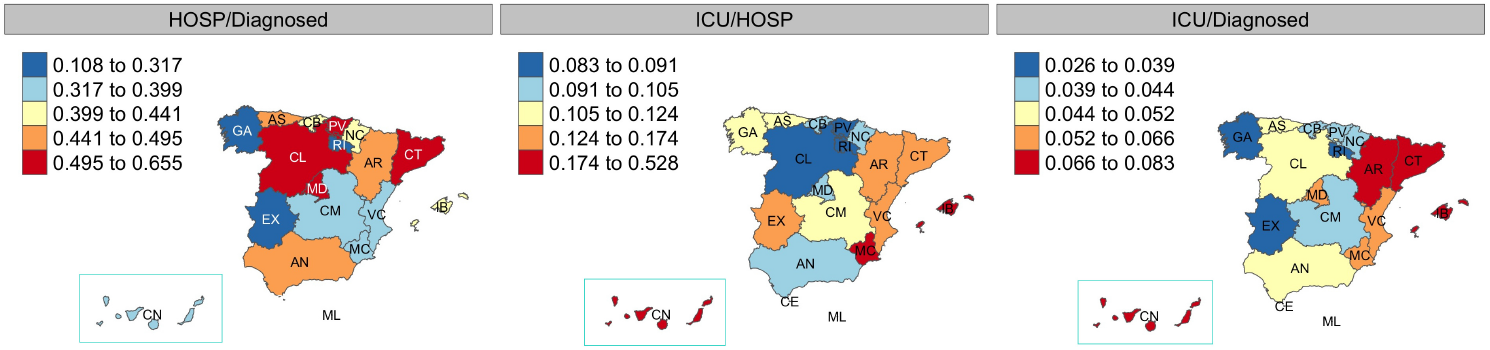
Mean of study variables by region: Proportion of Hospitalized over Cases (left); ICU over Hospitalized (middle); ICU over Cases (right).

Figure 4 considers the time evolution of the three proportion variables per region. The diversity in the recorded patterns is striking. With regard to the ratio of Hospitalized over Cases (red line), some regions present a relatively constant ratio, such as Andalusia (AN), Aragon (AR) or Navarra (NC). Others show an increasing pattern, with saturation already achieved by 08-Apr, as is the case in Castilla-León (CL), Catalonia (CT), or Galicia (GA); or not yet, such as in Asturias (AS). Madrid (MD) and Castilla-la-Mancha (CM) had a strongly increasing initial phase, which transformed quite suddenly into a steadily decreasing phase. The ratio of ICU over Hospitalized (blue) and ICU over Cases (green) show less obviously striking variety, but can also be divided into roughly constant, increasing and (already) declining. We insist that these are ratios, and therefore do not directly reflect whether the absolute numbers are increasing more or less rapidly.

**Figure 4:**
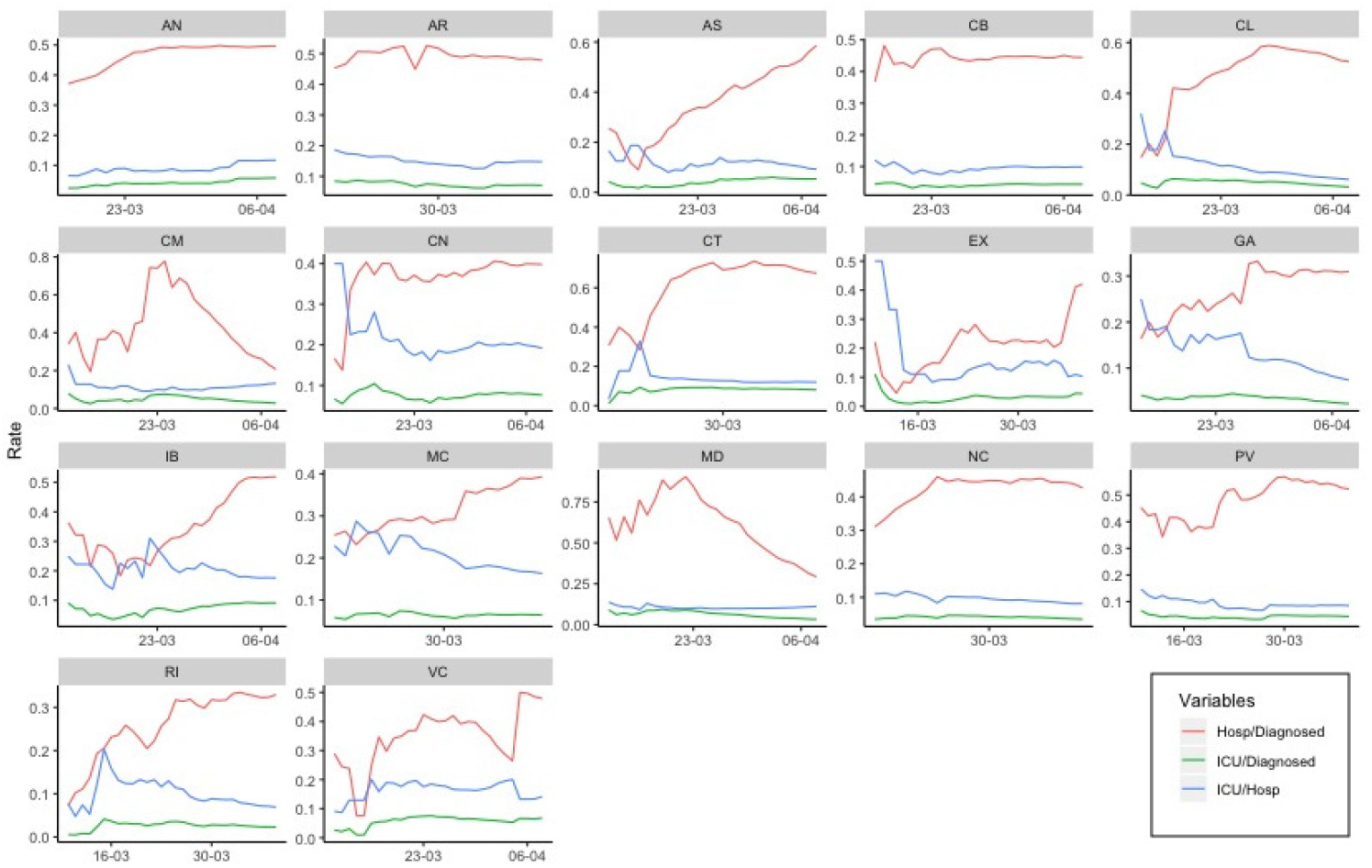
Evolution of study variables by Spanish region.

In the remainder of this paper, we will use the data up to 8-Apr to construct a model and make predictions, and verify this with the data from 9-Apr to 21-Apr, which is also provided by the Ministerio de Sanidad (2020). It should be noted that around 14-Apr the criteria for a patient to be counted as a “case” has changed in several regions, which started including asymptomatic patients from this date on. As stated before, the accuracy of the predictions could be further improved by introducing more reliable data. But it is worth stressing that the model itself does not depend on this, and the predictions are reliable and accurate within the assumptions and limitations of each counting method.

## 3 Methodology

This paper uses a combination of statistical and econometric analysis in different steps. First, a Beta distribution is used to estimate the density and distribution function of the study variables (the ratios defined in Eqs. 1) for both Spain as a whole and the Spanish regions separately. Note that this distribution does not involve any time parameter. Second, a logistic distribution is used to describe the time evolution of the cumulative raw data; in other words, the absolute number of diagnosed, hospitalized, and ICU patients, as well as recoveries, is estimated through the use of a logistic functional form. These auxiliary results are used as input for the third and main step, namely the Beta regression in which the ratios and their evolution is predicted, and which also includes an analysis of the temporal patterns and spatial effects in the variance of the modelization.

We begin by briefly summarizing these different elements.

### 3.1 Beta distribution

The Beta distribution *B*(*α, β*) is a family of two-parametric probability distributions limited to the interval *y* ∈ [0, 1]. It therefore naturally applies to probabilities and proportions, such as the ratio of hospitalized over total cases, or of ICU over hospitalized cases, that we are interested here. It is historically accredited to Karl Pearson, based on the Beta integral studied by Euler and Legendre in the 18th century.

The Beta distribution is characterized by a probability density function

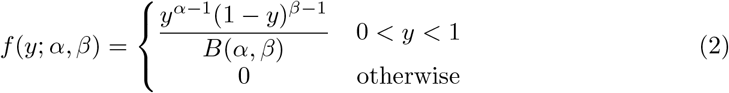

where the parameters *α; β* > 0, and the normalization constant

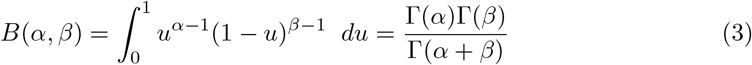

is precisely the Beta integral mentioned above, with Γ(·) the Gamma function.

It can be shown that the Beta distribution’s expected value or mean *µ* and variance *σ*^2^ are respectively.

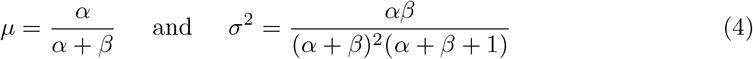

These parameters allow an alternative definition of the Beta distribution [Ferrari & Cribari-Neto (2004)] in terms of *µ* and a new parameter *ϕ*= *α* + *β*, such that *B*(*µ, ϕ*) is described by the equivalent probability density function

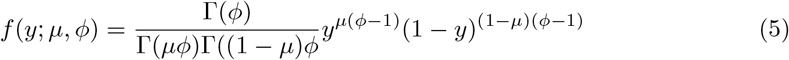

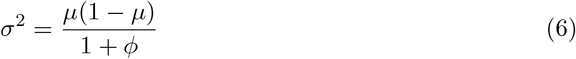

and therefore, for given mean *µ*, larger *ϕ*implies smaller variance. The parameter *ϕ*is therefore usually referred to as a precision parameter. This alternative parametrization forms the basis for the Beta regression model discussed below. We insist that this distribution describes the probability distribution of all data points, but does not contain an explicit time variable.

### 3.2 Logistic growth model

Logistic distributions form the basis for many population models in general [Richards (1959], including epidemic outbreak models [Chowell (2017)], and have been applied to the COVID-19 pandemic for example in Wu et al. (2020).

One possible way of writing a logistic population model is

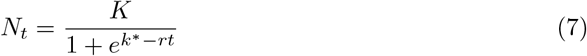

where *N*_*t*_ represents the population at time *t, k*^∗^ = *log*(*K − N*_0_)*/N*_0_, with *N*_0_ the initial population, *r* the growth rate, and *K* the carrying capacity (or asymptote). This is implemented in R through the parametrization Asym = *K*, scal = 1*/r* and xmid = *k*^∗^*/r*, see the table in Fig. 6a below. Note that Eq. (7) is obtained as a solution of the logistic or Verhulst equation, which is a first-order differential equation of the form

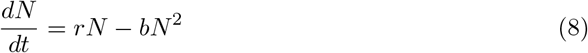

which describes a population (e.g., number of infected) whose size *N* grows in time proportionally with the existing population size, and with a quadratic saturation effect. This saturation can be due to competition for resources (food, space etc.) for a general population model, or to growing immunity, decrease of the non-infected population, or sanitary counter-measures in the case of an epidemic. We describe this in more detail in Appendix A.

### 3.3 Beta Regression

Since the dependent variables represent ratios or proportions, the final step consists of a Beta regression [Cribari-Neto & Zeileis (2009)]. Thus, for a random sample *Y* = (*y*_1_,, .., *y*_*n*_) of proportions, where it is assumed that each *y*_*i*_ ∼ *B*(*µ*_*i*_, *ϕ*_*i*_), the Beta regression model consists of two predictor functions. First, for the means *µ*_*i*_:

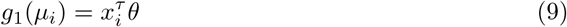

where *θ* = (*θ*_1_, …, *θ*_*k*_)^*τ*^ and *x*_*i*_ = (*x*_*i*1_, …, *x*_*ik*_)^*τ*^ are *k ×* 1 vectors representing the regression parameters *θ*_*j*_ and the independent variables (or covariates) *x*_*ij*_, respectively, and *g*(.) is a so-called link function. The independent variables *x*_*ij*_ could include pandemic features such as the lagged values of the same variables, as well as exogenous factors such as the proportion of recovered patients. To avoid problems of multicollinearity between variables, the correlation matrix and Variance Inflation Factors (VIFs) are calculated, see section 4.3. The vector of regression parameters *θ* (as well as *γ*, see below) is determined through a numerical maximum likelihood procedure based on the sample *Y*. The scalar product

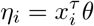

thus gives a linear prediction for the values of *g*(*µ*_*i*_). In models with a non-zero intercept (such as we will encounter here), the *x*_*i*1_ can be taken *x*_*i*1_ = 1 (∀*i*). The link function *g*(·) maps the restricted domain of the Beta distribution (*y* ∈ [0, 1]) to the range of the independent variables (in the most general case, ℝ), and can in principle be chosen freely in order to provide an optimal fit. A usual choice, which is known to provide good results in a wide range of applications and moreover is conceptually indicated for the current model because of its relation to the logistic function, is the Logit function.

A second predictor function is developed for the precision parameters *ϕ*_*i*_:

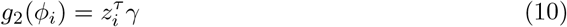

where *γ* = (*γ*_1_, …, *γ*_*p*_)^*τ*^ is the vector of regression coefficients and *z*_*i*_ = (*z*_*i*1_, …, *z*_1*p*_)^*τ*^ the set of independent variables. Note that basic Beta regression models use a constant parameter *ϕ*. However, we are interested in analysing the influence of temporal and spatial patterns, as well as cross effects, on the variance of the model, and these can ideally be studied by allowing for space and time-dependent precision parameters *ϕ*_*i*_, see Eq. (6).

In order to select which effects should be included in determining the *ϕ*_*i*_, an Akaike (AIC) and Bayesian Information Criteria (BIC) are applied [Akaike (1973) and Schwarz (1978)]. In these criteria, the following quantities

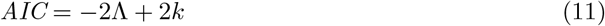

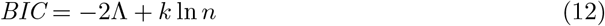

are minimized, respectively, where Λ represents the maximum log-likelihood, *n* the sample size and *k* the number of parameters (spatial, temporal or mixed precision coefficients) to be estimated by the model. These criteria thus incorporate a penalty for overfitting, ensuring that only a limited number of relevant parameters are selected.

## 4 Results

To illustrate the previously developed methodology, it will now be applied to the early phase of the COVID-19 epidemic in Spain.

### 4.1 Beta distribution analysis

We start by modelling the distribution of the dependent study variables defined in Eqs. (1) in Spain according to a Beta distribution. Table 1 shows the Beta distribution coefficients (*α*; *β*) from Eqs. (2) and (3) for the three ratios *HOSP/CASES, ICU/HOSP* and *ICU/CASES*. Note that all the obtained coefficients are extremely significant.

**Table 1:** Beta distribution: Parameter estimation

| Variable | Coefficient | Estimate | Std. Error | z value | Pr(>\|z\|) |
| --- | --- | --- | --- | --- | --- |
| HOSP/CASES | $\alpha$ | 3.490 | 0.223 | 15.67 | < 0.001 |
| | $\beta$ | 5.300 | 0.347 | 15.29 | < 0.001 |
| ICU/HOSP | $\alpha$ | 4.489 | 0.287 | 15.62 | < 0.001 |
| | $\beta$ | 27.324 | 1.833 | 14.90 | < 0.001 |
| ICU/CASES | $\alpha$ | 5.183 | 0.333 | 15.56 | < 0.001 |
| | $\beta$ | 96.760 | 6.514 | 14.85 | < 0.001 |

Figure 5 represents the density and cumulative distribution functions obtained from the parameters shown in Table 1 for the three ratios. The large difference between them is obvious from this figure. In particular, *ICU/HOSP* has a strong slope and faster cumulative progress in its probability distribution in comparison with the other two variables. This means that, according to the Beta distribution model, all the values for this ratio *ICU/HOSP* lie in the narrow range (0 − 0.15), whereas the other two ratios have a wider range.

**Figure 5:**
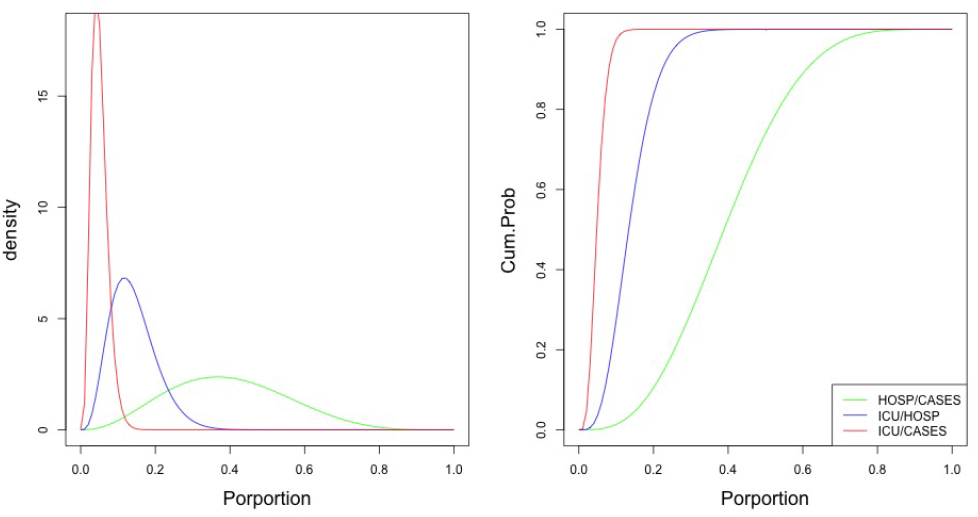
Beta density (left) and cumulative distribution function (right) for the results obtained in Table 1.

The graphical results of a statistical analysis showing that this Beta distribution model fits the empirical data to excellent degree is given in Appendix B. The density and distribution function of the study variables for each Spanish region separately are shown in Appendix C

### 4.2 Logistic prediction

The table in Fig. 6a records the estimated parameters of the logistic coefficients for the (absolute) cumulative number of COVID-19 cases, hospitalized patients, ICU patients, and recovered patients. All coefficients are highly significant, and a functional form is thus obtained to develop an estimation. Figure 6b plots these functional forms, as well as the real data and a forecast for the next two weeks. Since the fit between the functional forms and the real data is excellent, the forecast can be expected to be quite reliable, and this will be used to propagate the Beta regression and make forecasts for the ratio variables defined in Eq. (1). Nevertheless, it should be stressed that the logistic prediction is based on the simple logistic model that we have described in Section 3.2 and calculated for each variable separately. This gives a rather smooth prediction, with the drawback that any sudden variation in, for example, the number of hospitalized patients will not be accomodated by the next-day forecast for the number of ICU patients. In that sense, the Beta regression that we will describe next is more powerful, since it incorporates cross effects between the variables.

**Figure 6:**
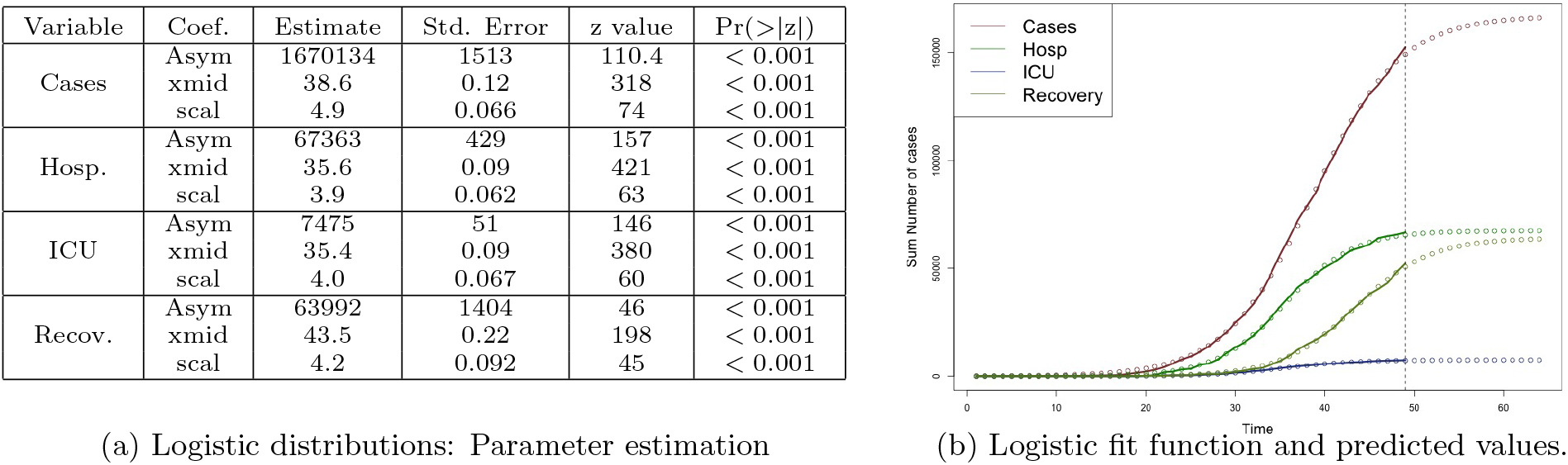
Logistic model and prediction. In (b), the dots represent real data values (left) and predicted values (right). The full line is the functional form obtained from the parameters of (a).

Logistic descriptions and predictions can also be made for each Spanish region separately. These are developed in Appendix D.

### 4.3 Regression model

In this section, a Beta regression model is developed for the ratios *HOSP/CASES, ICU/HOSP* and *ICU/CASES* defined in Eqs. (1a), (1b) and (1c). As independent variables, we consider the lags of the two first variables (i.e., their values one day before) as well as the recovery ratio (recovered patients over number of infections). The modelization also includes spatial patterns, temporal trends, and their cross effect through the precision coefficients.

To confirm that a meaningful regression model can be built with the independent variables just described, a twofold consistency analysis is performed previously. First, it is verified that the correlations between the independent variables are weak or at most moderate (|*R*| < 0.5). Second, a multicollinearity analysis shows that the VIFs or Variance Inflation Factors are low (*VIF* < 5) in all models, so there is no risk of meaningless overfitting [Fox & Monette (1992)]. For this reason, the model does not include the lag of the third ratio, *ICU/CASES*, since–as mentioned above–this can be obtained as the product between *HOSP/CASES* and *ICU/HOSP*, and it is thus easy to understand that it is strongly correlated with both, and thus leads to multicollinearity and high VIF problems. Further details are given in Appendix E.

A crucial element of the model is the precision parameter *ϕ*, defined in Eq. (6) and discussed in Section 3.3. This precision parameter determines the variance of the Beta distribution, and thus the reliability of the forecast. It can be taken either as a global parameter (a single *ϕ*for the whole distribution), as dependent on temporal effects only (*ϕ*evolves with time from the onset of the epidemics), as dependent on spatial effects only (a separate *ϕ*_*i*_ for each region *i*), or a mixed model which includes both temporal and spatial effects. Both the AIC and the BIC criterion indicate that in two of the three cases the mixed model is by far the best, in spite of the fact that it has the highest number of parameters (and thus is the most penalized), i.e.: both spatial and temporal effects are sufficiently relevant and should be taken into account. In the third case (ICU/Hosp), the mixed-effect model is slightly outperformed by the spatial-effects-only model, but since the difference is tiny we also apply a mixed-effect model here. Details of the AIC and BIC are given in Appendix F.

Table 2 shows the coefficients obtained for the Beta regression model of the study variables. The precision coefficients are given in Appendix C. The very high significance levels and Pseudo-*r*^2^ values indicate that the obtained Beta regressions provide excellent models for the empirical data. This is confirmed by the application of the model to the data for Spain as a whole, as shown in Fig. 7, as well as for the regions separately, see Appendix G. The incidence of the factors considered is quite different. For the Hosp/Cases ratio, the lagged value of Hosp/Cases itself is much more important than the lagged value of ICU/Hosp, and vice versa, which seems reasonable. For the third ratio, ICU/Cases, the contribution of both lagged values is of the same order of magnitude. The recovery rate, finally, has a small but statistically significant influence on the various ratios. Curiously, this influence is in fact positive on the ICU/Hosp ratio: an increment in the recovery rate leads to a (small) increase in the ICU/Hosp ratio. This can probably be understood as follows: patients who are considered “recovered” in the early phase of the epidemic are likely to come from the “light” (non-ICU) category of patients. Removing them from the pool of Hospital patients therefore automatically increases the relative share of severe (ICU) cases.

**Table 2:** Beta Regression Results

| Hosp/Cases |  |  |  |  |
| --- | --- | --- | --- | --- |
|  | Estimate | Std. Error | z value | Pr(>\|z\|) |
| Intercept | -1.258 | < 0.001 | -12001 | < 0.001 |
| Lag Hosp/Cases | 2.515 | < 0.001 | 11567 | < 0.001 |
| Lag ICU/Hosp | 0.001 | < 0.001 | 4.03 | < 0.001 |
| Recovered Rate | -0.001 | < 0.001 | -11.1 | < 0.001 |
| ICU/Hosp |  |  |  |  |
|  | Estimate | Std. Error | z value | Pr(>\|z\|) |
| Intercept | -1.792 | 0.001 | -1358 | < 0.001 |
| Lag Hosp/Cases | 0.016 | 0.001 | 13.6 | < 0.001 |
| Lag ICU/Hosp | 5.012 | 0.016 | 323 | < 0.001 |
| Recovered Rate | 0.012 | 0.001 | 14.9 | < 0.001 |
| ICU/Cases |  |  |  |  |
|  | Estimate | Std. Error | z value | Pr(>\|z\|) |
| Intercept | -2.596 | 0.005 | -531 | < 0.001 |
| Lag Hosp/Cases | 1.159 | 0.009 | 125 | < 0.001 |
| Lag ICU/Hosp | 3.718 | 0.023 | 164 | < 0.001 |
| Recovered Rate | -0.087 | 0.008 | -11.2 | < 0.001 |

**Figure 7:**
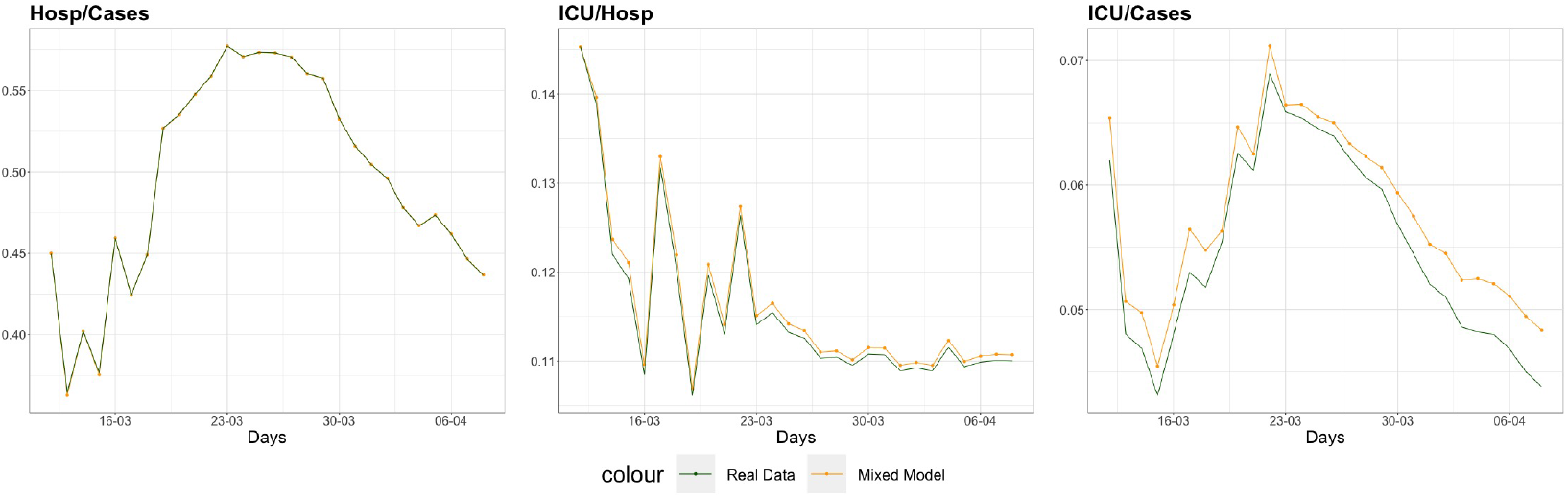
Beta regression model for Spain until 8-Apr for the ratios Hosp/Cases (left), ICU/Hosp (middle) and ICU/Cases (right)

The complete list of the precision coefficients *ϕ*_*i*_ is given in Appendix H. Here, we limit ourselves to some general observations. The spatial patterns are highly significant for a majority of the Spanish regions, for all three ratios. The temporal trends are highly significant for Hosp/Cases and ICU/Hosp, but not for ICU/Cases. The cross effects (spatial and time) are, again for all three ratios, highly significant for a majority of the Spanish regions.

With respect to their concrete incidence, it is striking that the cross effects for each single region are negative with respect to Hosp/Cases and positive (with one single exception) with respect to ICU/Cases. We don’t have a straightforward interpretation for this phenomenon, but the main conclusion is that space-time cross effects are relevant for the determination of the precision parameter, and hence for the Beta regression model in general and its dispersion in particular.

Figure 7 plots the mixed Beta regression models for the three ratios for the period until 08-Apr. This mixed model (with space-time cross effects) is seen to adjust very closely to the real data, and so will be used next to make forecasts.

### 4.4 Forecast

Figure 8 represents the next-day prediction for the three ratios according to the Beta regression results. The horizontal plane of this 3D plot represents all possible lagged values of the Hosp/Cases and ICU/Hosp ratios (on a scale from 0 to 1), i.e.: the previous values of these variables (at a lag of 1 day). The vertical axis then shows the Beta regression prediction for the dependent variable 1 day later, from left to right: Hosp/Cases, ICU/Hosp and ICU/Cases. In this prediction, for graphical convenience, a single constant recovery rate is assumed, namely 8%, which is the mean recorded by the study database. Obviously, this prediction can be further improved numerically by adjusting this recovery rate.

**Figure 8:**
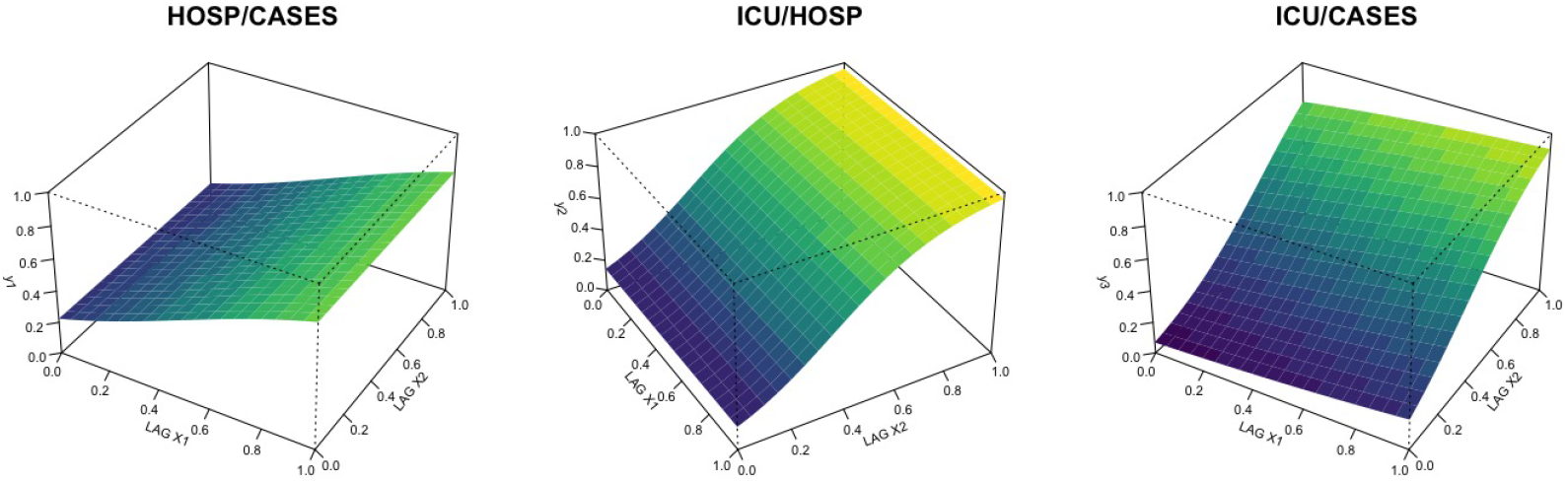
Prediction by of Hosp/Cases, ICU/Hosp and ICU/Cases for a recovery rate of 0.08. The independent variables are the 1-day lagged values, in particular X1 the lagged ratio Hosp/Cases, and X2 ICU/Hosp.

Once this has been established, the main point of interest is to make predictions for the real data for each ratio. This is represented in Fig. 9. Two predictions are plotted here for the period 9-Apr to 21-Apr: the next-day predictions based on the real data, and a prediction based on the logistic forecast of the number of diagnosed, hospitalized, ICU patients and recovered cases. In other words, to calculate the prediction for, for instance, 15-Apr, the real-data next-day prediction is based on the Beta regression prediction obtained from the recorded ratios 14-Apr, whereas the logistic forecast uses the recorded data only up to 8-Apr, propagates this into a logistic forecast up to 14-Apr, which is then fed into the Beta regression to obtain a prediction for the ratios on 15-Apr.

**Figure 9:**
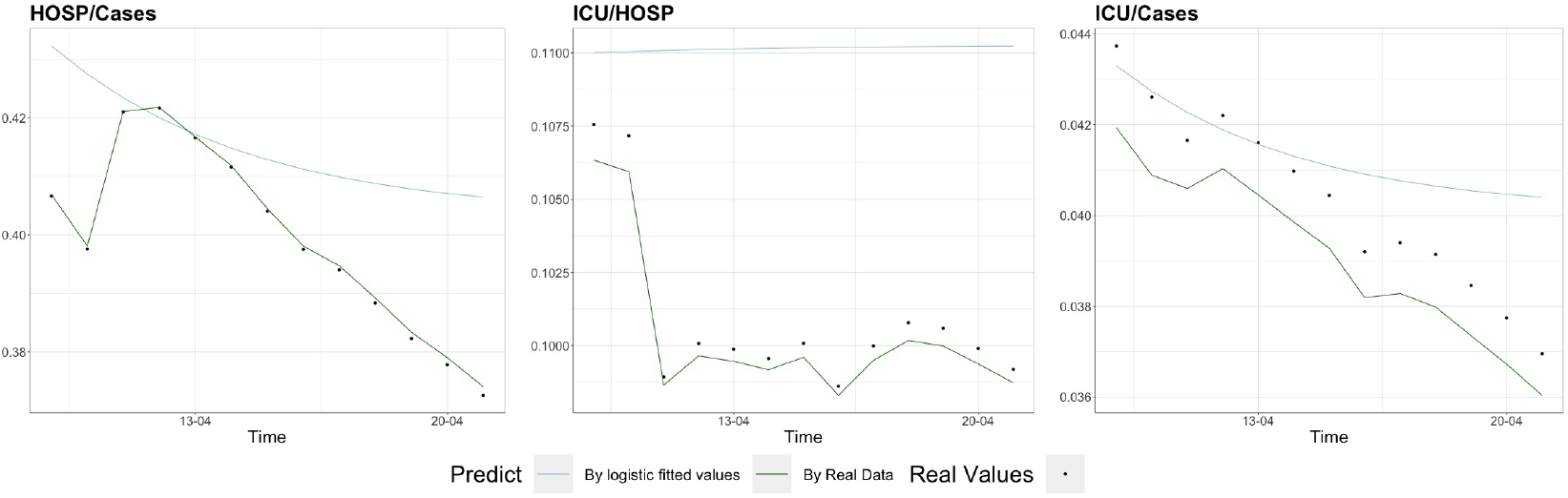
Prediction of the three ratios Hosp/Cases, ICU/Hosp and ICU/Cases for Spain as a whole based on the Beta regression and comparison with real data for the period 9-Apr to 21-Apr. In all three figures, the Y-axis range is strongly adjusted to the range of values.

As can be seen, the next-day predictions are extremely close to the real data. The multiple-day prediction obtained through the fitted values from the logistic expression of Table 6a shows a stronger difference with respect to the real values. However, both in relative and in absolute terms this difference is still very reasonable (or said in other words, the scale of the graphs is strongly adjusted to the range of values, which might lead one to visually overestimate the error).

Nevertheless, it is worth analysing the origin of these differences between the logistic-based Beta regression and the real data. First, there is the intrinsic fact that this forecast is used for a prediction several days ahead, whereas the real-data prediction uses the empirical data from the day before and so is much more cautious.

However, the main reason for the differences lies in the poor quality of the data. Two issues stand out here. First, the different regions have used different criteria and methods to count the number of cases. Second, these criteria have even been changed precisely in the period that we use to make our prediction, which is particularly obvious from the jump in Fig 8 in the first few days of the prediction. Obviously, this makes it highly complicated to make any accurate predictions, and makes it very difficult for hospital managers to make any reasonable planning. The simple logistic model that we have used (see also Appendix A) is particularly vulnerable to this, since it does not adapt from one day to the next, which also contributes to the relatively large errors shown in Fig 9. More advanced logistic models, see e.g. [Wu et al. (2020)] could be used to predict the absolute numbers of cases, and derive from it a Beta prediction for the ratios that we have studied.

Finally, it should be stressed that the logistic-based forecast gets the slope essentially correct, and in fact even ascertains the correct value within an error margin of roughly 10% over the complete time range of prediction performed here (i.e., 12 days). This illustrates the robustness and usefulness of the methodology.

The predictions are further specified by region in Appendix I.

## 5 Conclusion

Logistic models are the most commonly used statistical models for the study of epidemic evolution. In this article, we have discussed how these can be complemented through the use of Beta distribution and regression models. Beta models are useful for variables whose values are limited to the range [0, 1] and are thus naturally suited to describe proportions. In particular, we have described how they can be used to describe the ratio between hospitalized patients and diagnosed cases (Hosp/Cases), between ICU patients and hospitalized cases (ICU/Hosp), and between ICU patients and diagnosed cases (ICU/Cases). These variables are particularly relevant for hospital management in the early phase of an epidemic crisis such as the one that we are currently experiencing due to the COVID-19 virus. Indeed, these ratios are indicators of the strain put on hospital resources in general, and of ICU units in particular. Moreover, they have the advantage of being scale-independent, and therefore adaptable to any hospital or region of any population size. They can thus help in planning contingency measures, making resource estimates, and reserving hospital capacity across all hospitals in a given region or country, independently of size. They also provide a robust way of planning the distribution of temporary facilities dedicated exclusively to the epidemic. The Beta regression model developed here depends on the 1-day lagged value of the Hosp/Cases and ICU/Hosp ratios, as well as the patient recovery rate. The lagged value of the third ratio, ICU/Cases, was not used in the model as it would lead to dependency and multicollinearity issues. It was also shown that the most performant model includes mixed space-time effects in the precision coefficient of the Beta distribution. These precision coefficients are indicators of (inverse) variance, and allowing them to vary across regions and along time thus allows to minimize the resulting variance.

This model has been applied to the early-phase Covid-19 epidemic in Spain. It has been shown that next-day predictions for these ratios based on a Beta regression model are extremely reliable. For longer-term predictions, the error was obviously larger. The main cause of this error was pinpointed to lie in the lack of consistency and reliability of data in this early phase of the epidemic. This became obvious even at the Spanish level: different regions have used different criteria to count COVID-19 cases, possibly also of counting COVID-related hospitalized and ICU patients, and have even changed their criteria during the crisis. This renders highly difficult a serious statistical analysis of the situation, and more importantly: makes it highly difficult to make serious hospital management predictions and provisions. In this sense, universal guidelines on how to collect, classify and publish data for such pandemics, for example issued by the WHO, might be useful. Nevertheless, the methodology has been shown to be rather robust against these difficulties and to provide relatively reliable forecasts. In particular, for a 12-day prediction from 9-Apr to 21-Apr, the logistic-based Beta regression still gets the slope essentially correct, and the concrete values obtained for the three ratios that were analysed remain within an error margin of 10% compared to the real data.

Finally, it should be pointed out that the Spanish Ministry of Health has in the meantime issued updated data. We plan to apply the method developed here on these updated data sets. In particular it would be interesting to see whether propagating the Beta regression without the auxiliary logistic step leads to reliable and robust predictions of the ratios when applied to such updated and more realistic data.

## Data Availability

None

## Acknowledgments

G.J. acknowledges project FIS2017-86497-C2-1-P of the Spanish MICINN.

## Appendices

### A Logistic model: theoretical comments

In this Appendix, we briefly review basic mathematical population growth models and how these can be solved with differential equations, see e.g. [Boyce & DiPrima (2012)].

Imagine a population *N* (e.g., number of infected) which increases with time *t* at a rate *r* proportional to the existing population (existing number of infected):

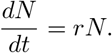

The solution is

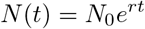

where *N*_0_ = *N* (*t*_0_) represents the initial population.

Taking the (natural) logarithm on both sizes, this can also be written as

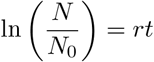

or, for any time difference:

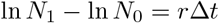

The main drawback of this model is that it does not include saturation effects. As a population grows larger, competition for resources (such as food) come into play. If the considered model regards a subpopulation (e.g., number of infected), the growth rate could also decline because of the corresponding decrease in the pool of non-infected population members, or because of sanitary counter-measures and forced social distancing. Such saturation effects can most simply be modelled as a negative quadratic influence, i.e.:

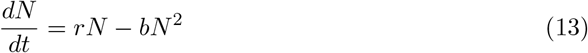

This is structurally identical to the well-known logistic equation, whose solution is therefore the logistic or Verhulst function, which in this case can be written as

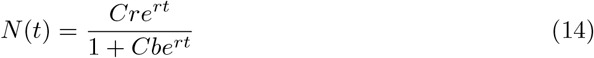

where 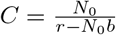 In the long term the population tends to the (stable) value

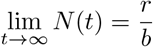

regardless of the initial population *N*_0_. By defining *K* = *r/b*, the expression given in Eq. (7) is found. The function (14) has the typical sigmoid (*S*-shaped) form seen in Fig. 6b. Finally, let us briefly look at the case in which one is interested in a proportion between two populations. Assume that both are described by a simple exponential evolution as described above

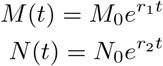

The ratio 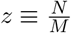 then follows

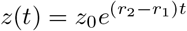

where *z*_0_ = *N*_0_*/M*_0_. Only if both populations grow at exactly the same rate (*r*_1_ = *r*_2_), a non-exponential result is found, namely the constant ratio

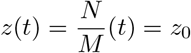

Likewise, it can be shown that the inclusion of saturation effects will still lead to a logistic function for the ratio of populations.

### B Beta Distribution for Spain: comparison of model with data

Figure 10 compares the Beta distribution obtained as a fit for the three ratios, Hosp/Cases, ICU/Hosp and ICU/Cases. In each case, the top-left plot provides a comparison between the functional form of the proposed models and the empirical cumulative distribution. The top-right graphs show the corresponding histograms. The two bottom figures represent the Q-Q and P-P plot, i.e. a comparison between theoretical and empirical values in terms of quantiles (left) and of probabilities (right). The agreement in all cases is excellent, with the exception of t

**Figure 10:**
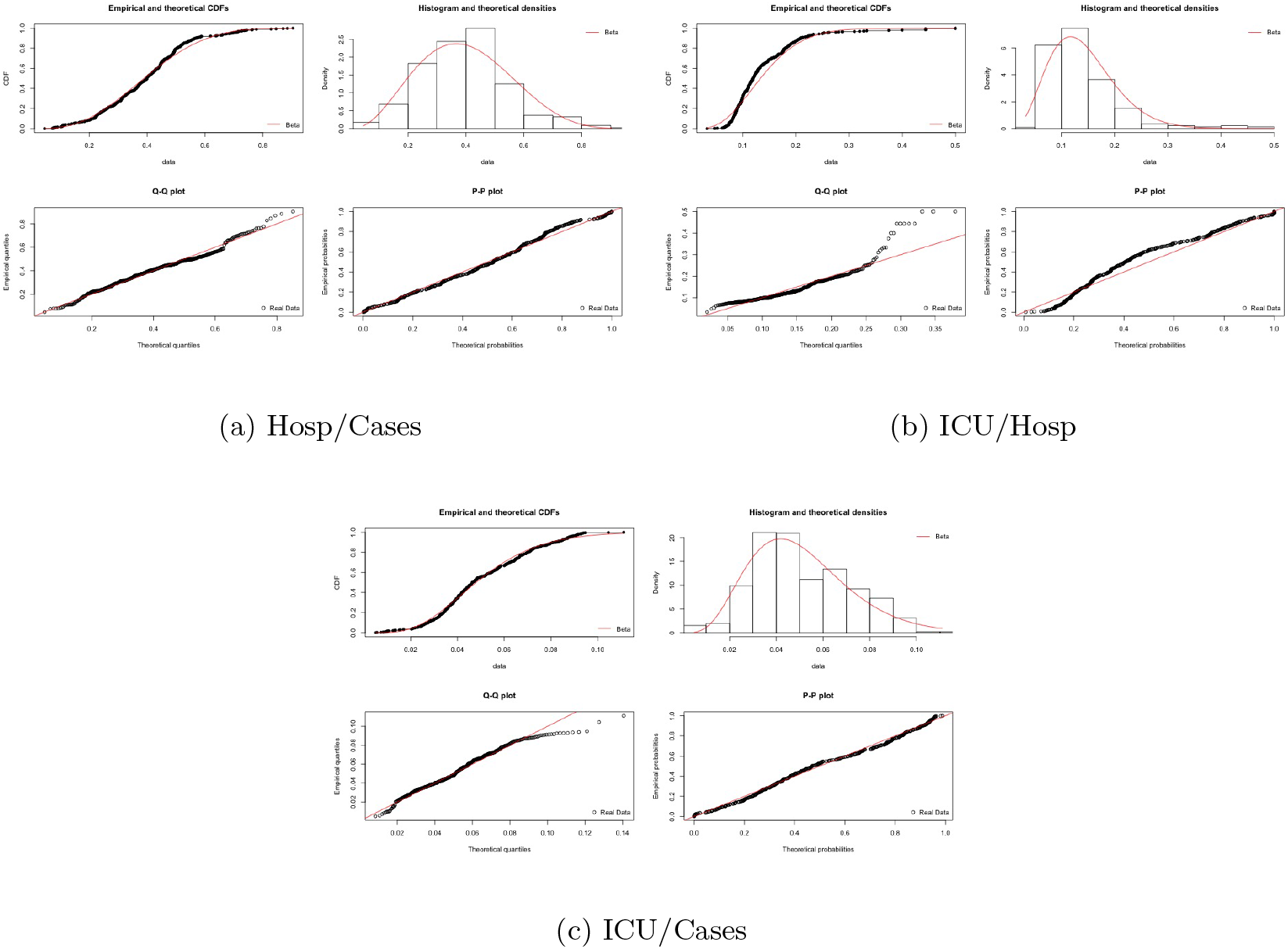
Comparison of Beta distribution models and empirical data. See text for details. In each case, the solid line represents the theoretical Beta distribution model, the dots (or bars, in the case of the histogram) the empirical data.

The apparent discrepance in the Q-Q plots at high ranges is an artifact of this quantile representation, as can be seen from the other graphs. All in all, the agreement between the models and the empirical data is excellent.

### C Beta Distribution per Spanish region

Section 4.1 contained the Beta distribution analysis for Spain as a whole. Fig. 11 similarly shows the Beta distribution and cumulative density function for each region separately. There is a relatively large variety of slopes and patterns. This can be seen for the ratio HOSP/Cases, which is more extended in Madrid, Catalonia, or Castilla-León than in other regions, and especially than in Aragon (AR), Murcia (MC) or Navarra (NC). It is also true for the ratio ICU/Hosp, which has a rather large spread in Extremadura (EX), Canary islands (CN) or Castilla-León (CL), and is much more peaked in Madrid (MA), Andalusia (AN) or Aragon (AR). The ratio ICU/Cases, finally, looks more homogeneous across the regions, with a sharp peak at low values.

**Figure 11:**
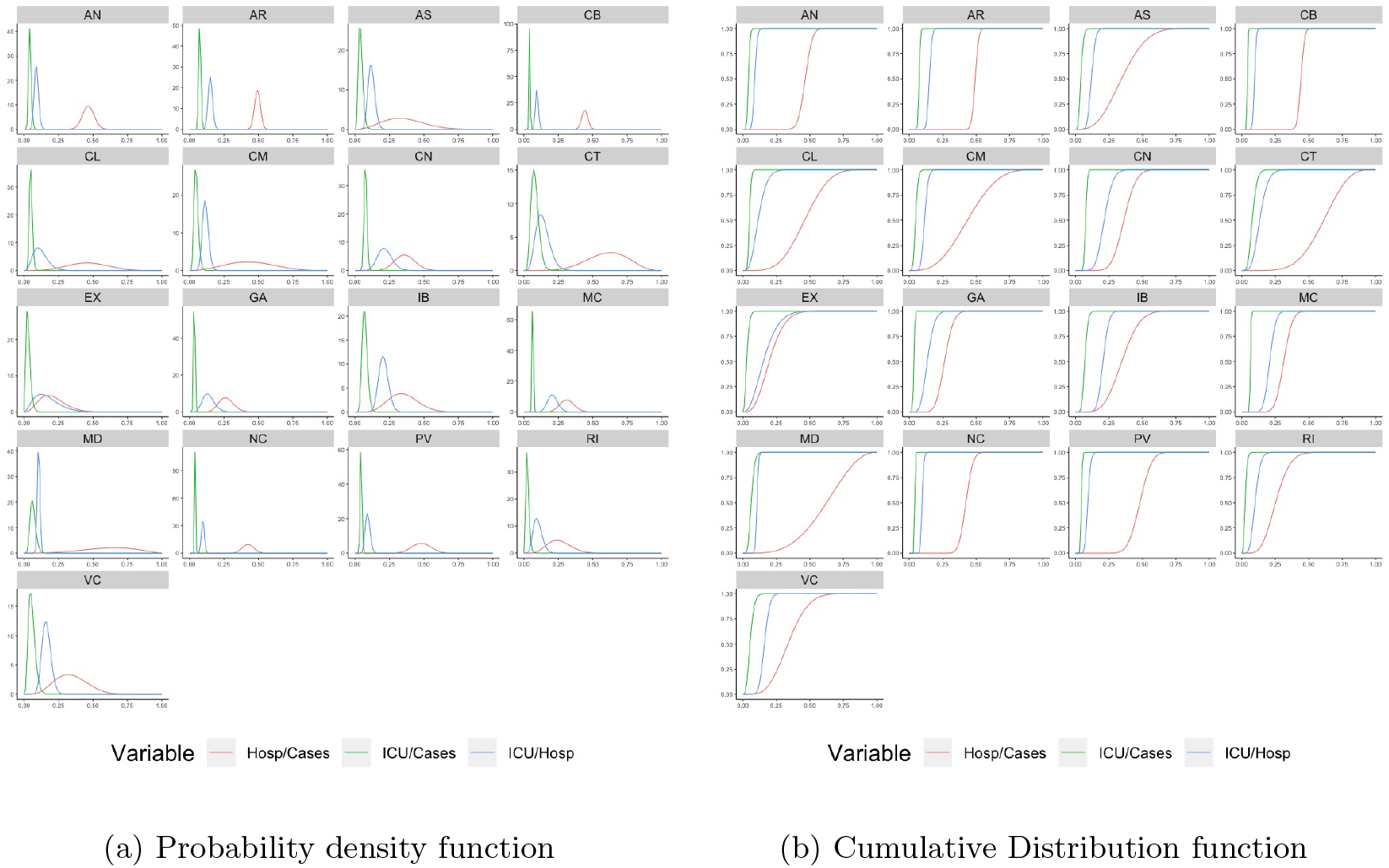
Beta density and cumulative distribution function per Spanish region

### D Logistic model per Spanish region

A logistic model and prediction was developed for the whole of Spain in Section 4.2. Here, the results for the logistic model for each Spanish region separately are plotted for the absolute (cumulative) number of cases in Figure 12a, of hospitalized patients in Figure 12b, ICU patients in Figure 12c and recovered patients in Figure 12d. In each figure, the red line represents the real data (as previously, obtained from [Ministerio de Salud (2020)] up to 8-Apr, and the blue line describes the values according obtained from the logistic functional form, region per region, which are extended to a prediction up to 21-Apr.

**Figure 12:**
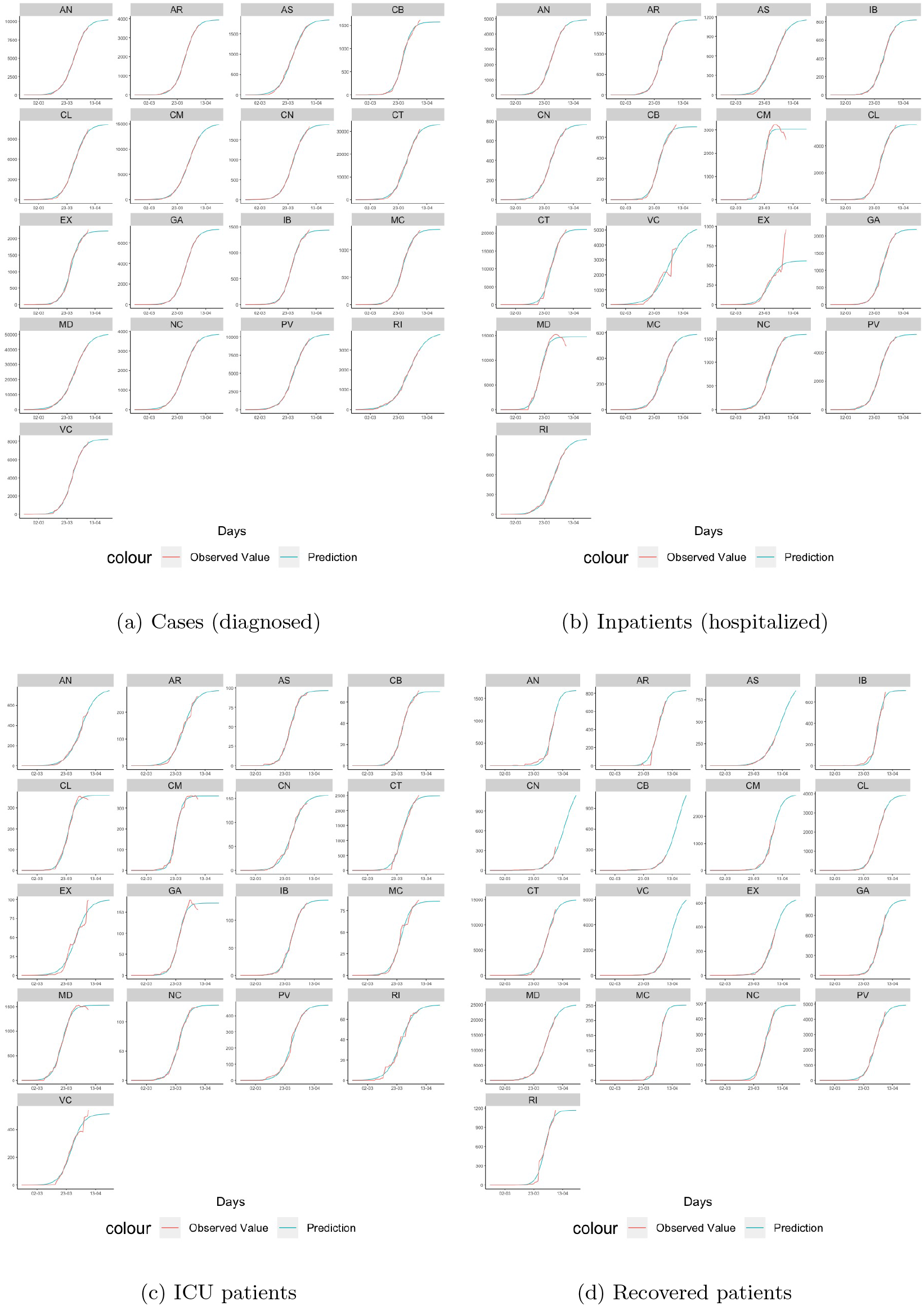
Logistic Analysis per Spanish region: absolute cumulative numbers of observed values as published by the Spanish Ministry of Health [Ministerio de Sanidad (2020)] versus logistic prediction

In most cases, the fit between the model and the real data is excellent, especially in the early period studied. In some regions, such as Extremadura (EX), Castilla-La Mancha (CM) or Valencia (VC), the empirical pattern changes rather abruptly towards the end of the recorded data, and this leads to larger differences between the model and the data. As mentioned in the main text, this is most likely due to a change in the counting methodology or classification criteria adopted by the regions during this period, which becomes particularly clear in the (temporary) decline in the empirical data in some of the regions, even though these represent cumulative numbers.

### E Correlation and multicollinearity analysis

When constructing a regression model, it is important to verify that the correlation between all possible pairs of variables are low, and that there are no multicollinearity problems. Otherwise, this could lead to meaningless overfitting and spurious results.

The correlation matrix in Fig. 13a shows that all pairs of variables show adequate correlation values (*R* < 0.8) and can therefore be used in the regression model.

**Figure 13:**
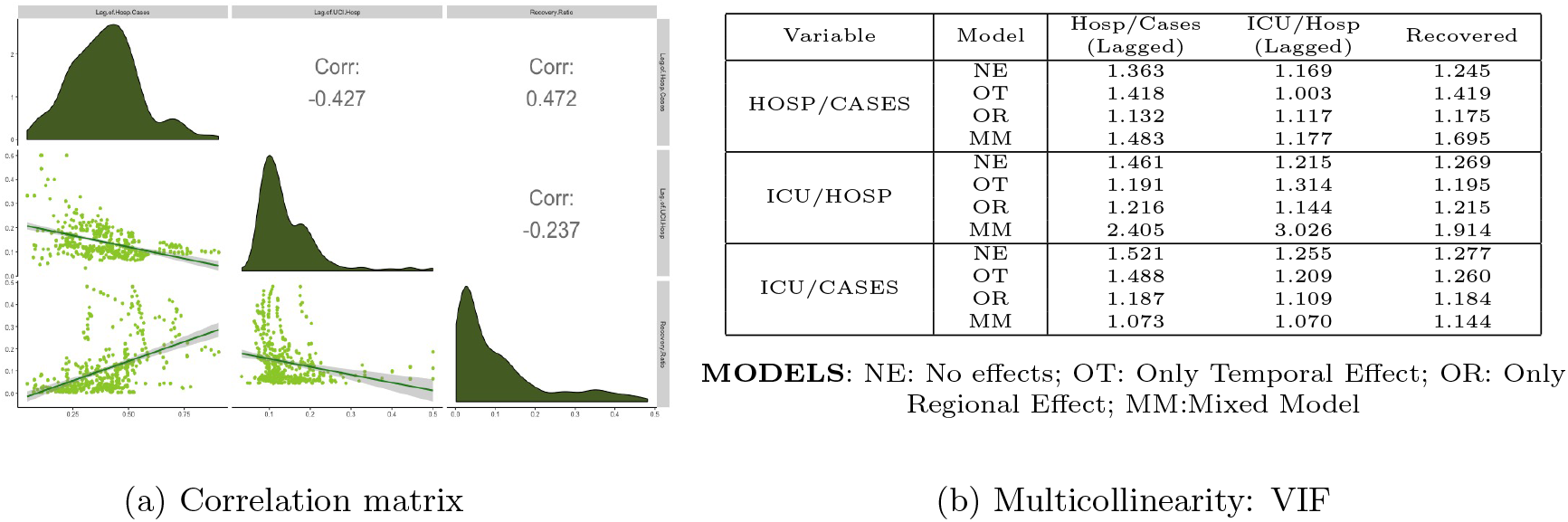
Correlation matrix and collinearity analysis of the variables used in the regression models.

The VIF (Variance Inflation Factor) for each model (no effects, temporal effects only, spatial effects only, mixed space-time effects) are shown in the table of Fig. 13b. Low values are obtained in every model (*VIF* < 5), so the models avoid the multicollinearity problem.

Note that the inclusion of the lagged value of ICU/Cases has also been considered, but since this presented high VIF values as well as strong correlation with the other variables, it has not been withheld as an independent variable.

### F Model Selection: Akaike and Bayesian Information Criteria

Table 3 shows the AIC and BIC values obtained for the different models (no effects, i.e. constand precision parameter *ϕ*; only spatial effects; only temporal effects; or mixed spacetime effects) when applied to the three ratios Hosp/Cases, ICU/Hosp and ICU/Cases. For both Hosp/Cases and ICU/Cases, the Mixed model has by far the lowest value according to both criteria. For ICU/Hosp, the mixed model actually is slightly outperformed by the model with only spatial effects, indicating that the inclusion of mixed effects leads to a slight overfitting. However, since the difference is very small, and for consistency with the other cases, a Mixed model has been selected in this case also.

**Table 3:** Regression model selection through AIC and BIC information criteria.

| Hosp/Cases | model | BIC | AIC |
| --- | --- | --- | --- |
|  | No Effect | -2508.635 | -2529.080 |
|  | Only Region effect | -3067.087 | -3152.957 |
|  | Only Temporal Effect | -2813.489 | -2838.024 |
|  | Mixed Effects | -3841.464 | -3996.848 |
| ICU/HOSP | model | BIC | AIC |
|  | No Effects | -2769.363 | -2789.964 |
|  | Only Region effect | -3091.766 | -3177.636 |
|  | Only Temporal Effect | -2947.351 | -2971.885 |
|  | Mixed Effects | -3069.672 | -3159.631 |
| ICU/Cases | model | BIC | AIC |
|  | No Effects | -2936.698 | -2957.143 |
|  | Only Region effect | -3159.396 | -3183.930 |
|  | Only Temporal Effect | -3160.522 | -3246.392 |
|  | Mixed Effects | -3435.505 | -3590.889 |

### G Beta regression model per Spanish region

Fig. G shows the Beta regression model using mixed space-time effects and a comparison with the empirical data for each ratio and for each Spanish region separately. It can be seen that there is a wide diversity of patterns across the regions. However, the Beta model adjusts to all of them, in most cases to an excellent degree.

### H Estimation of Beta Precision Coefficients

Because of the results of the AIC and BIC analysis, see Appendix F, the Beta regression model assumes that the precision parameter is not constant, but depends both on the evolution of time and on the concrete Spanish region, as well as on space-time cross effects. These precision coefficients by time and region, as well as their cross effects, are listed in table 4.

**Table 4:**
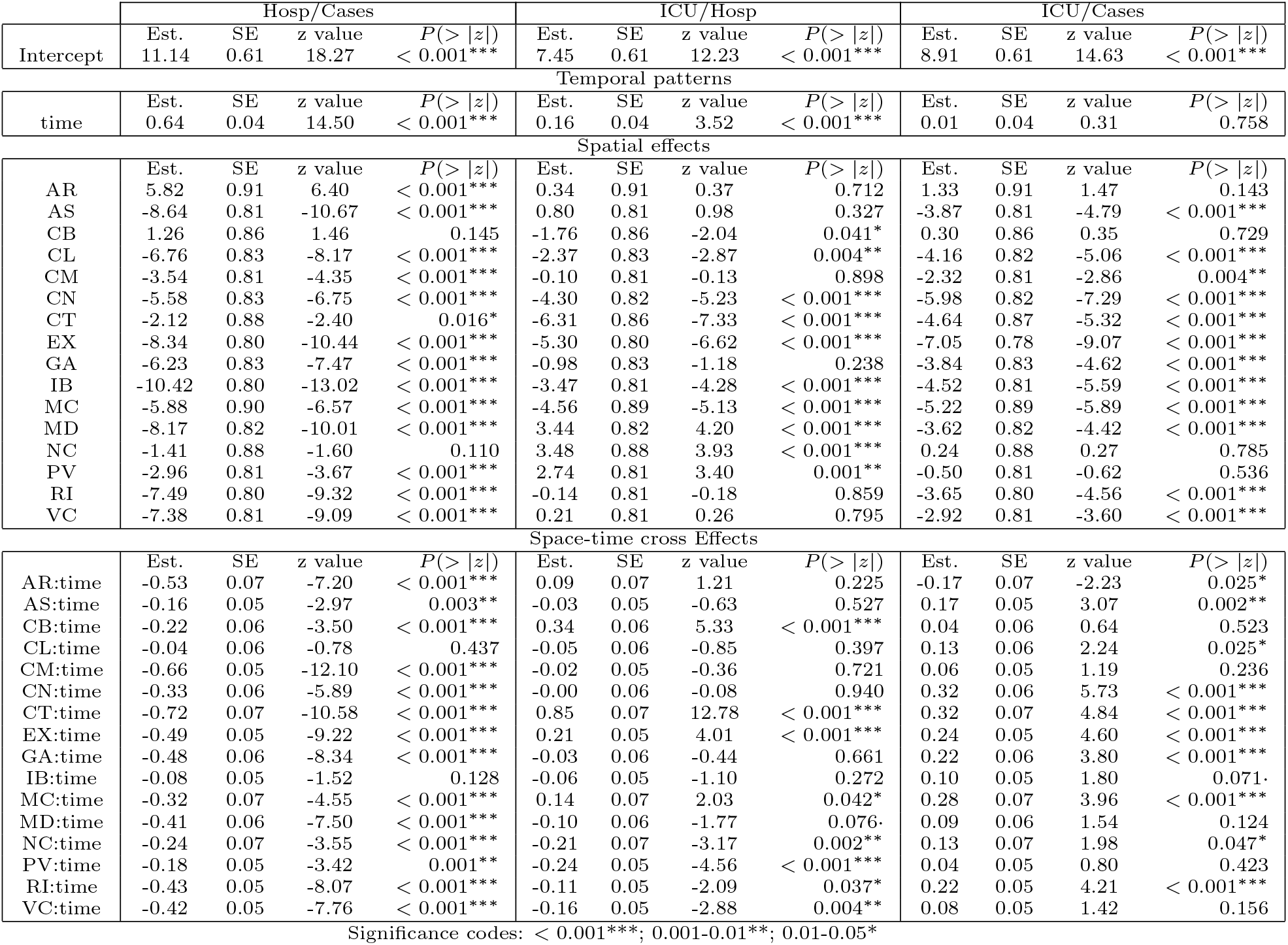
Precision coefficients according to each Beta regression

Note that most coefficients are significant only for two (or exceptionally even only one) of the ratios. However, overall, each block is highly significant, at least for a majority of the regions. This is in agreement with the result of the AIC and BIC analysis just mentioned. To illustrate this, when looking at the temporal patterns, these are not significant for ICU/Cases, but they are highly significant for both Hosp/Cases and ICU/Hosp. An analogous observation applies to the spatial coefficients: for most regions, the spatial coefficient is not significant for one of the ratios – for Aragon (AR), Cantabria (CB) and Navarra (NC) even for two of the ratios. However, they are highly significant for the two other ratios – or the single remaining ratio for the three regions just mentioned. The significance of the cross effects could be detailed in a similar way.

### I Prediction by Spanish region

Fig. 15 shows forecasts per Spanish region for the three ratios Hosp/Cases, ICU/Hosp and ICU/Cases. It shows similar strengths and weaknesses as Fig. 9 for Spain as a whole. The next-day prediction based purely on the Beta regression using the 1-day lagged ratios is excellent in almost all cases.

**Figure 14:**
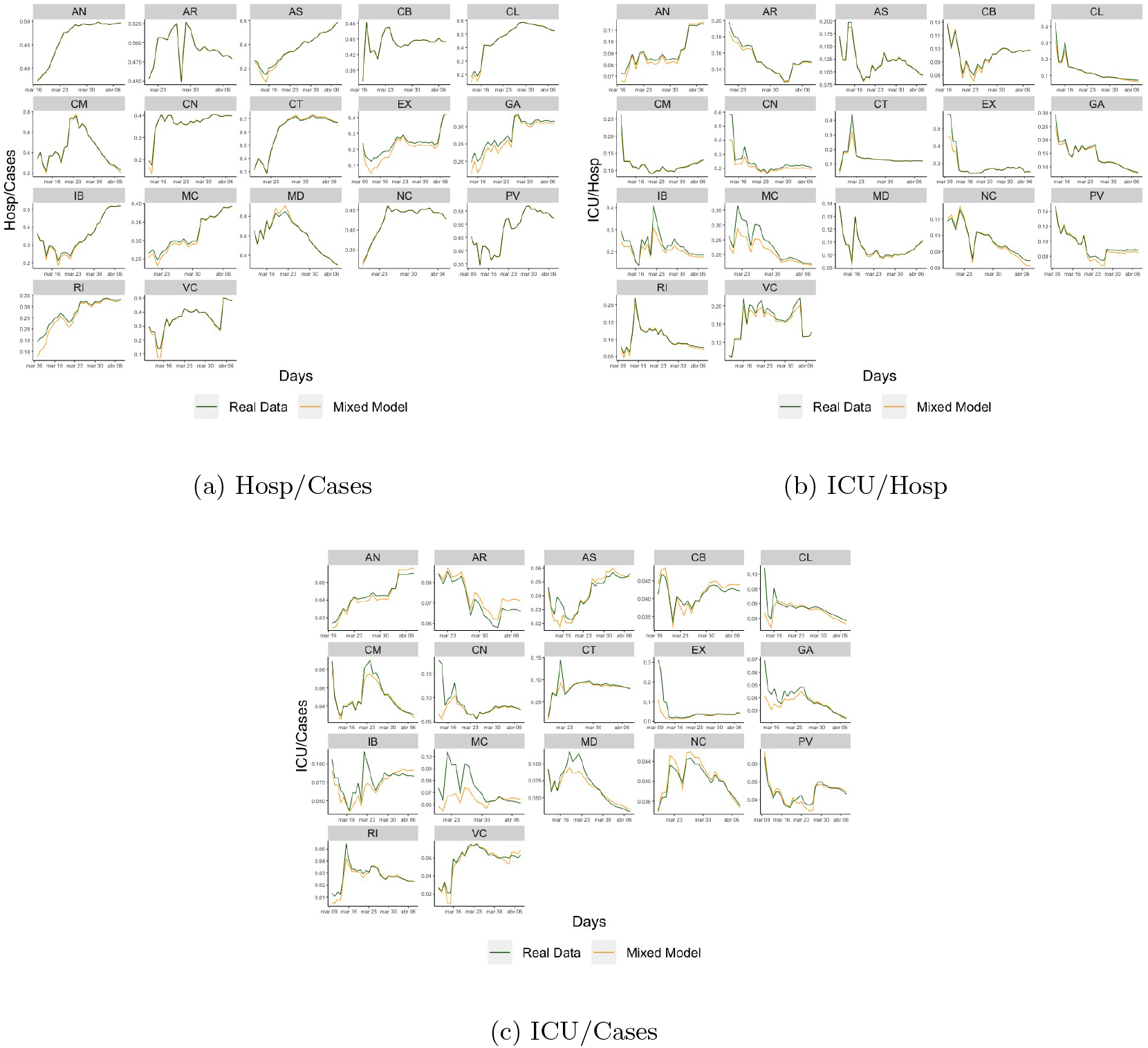
Beta regression model and comparison with empirical data per Spanish region

**Figure 15:**
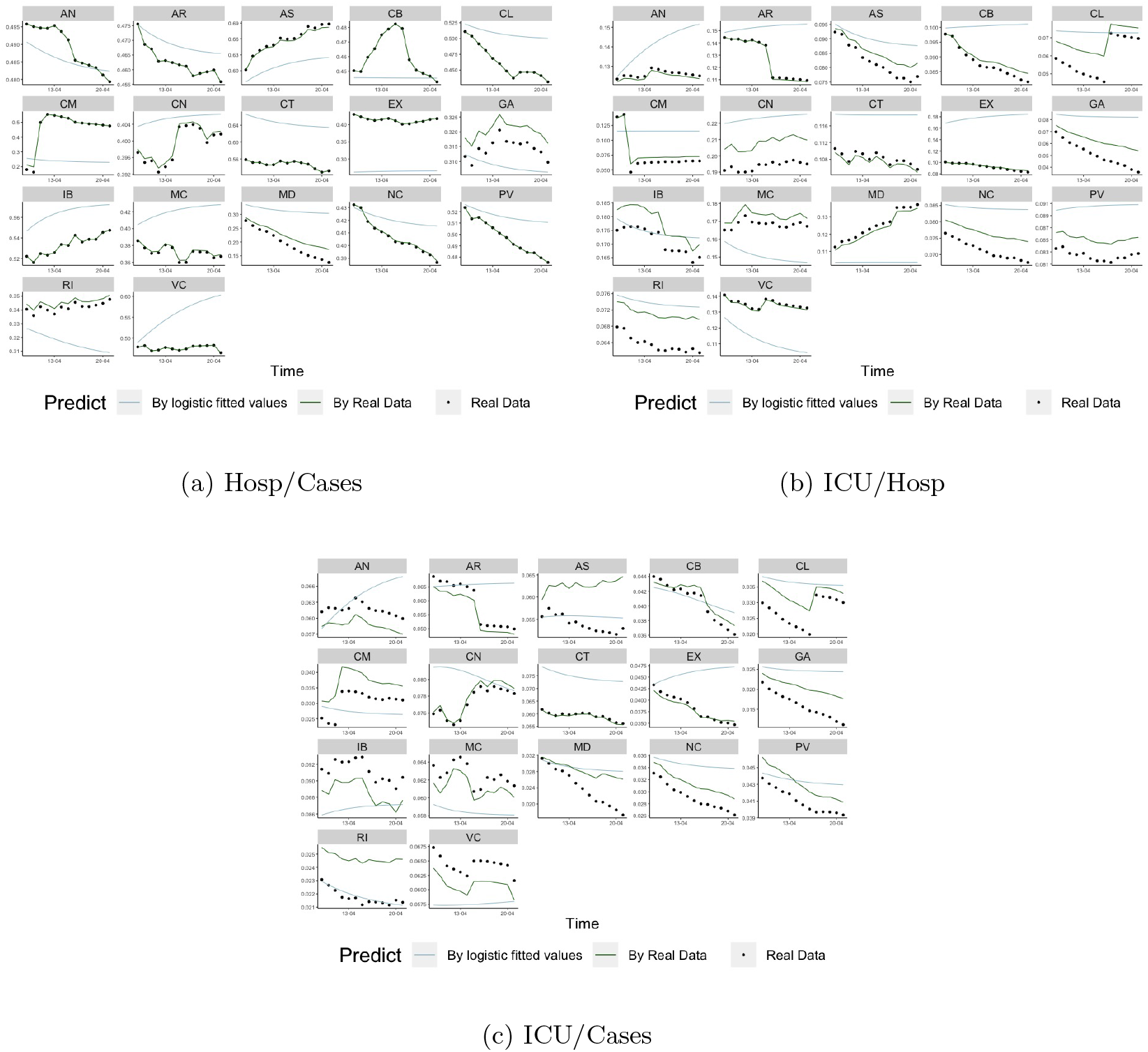
Prediction by Spanish region using real data and using logistic propagation model

The multiple-day prediction based on the logistic forecast is quite good for most variables in most regions, although there are some cases with a relatively wide difference.

## Notes

### Competing Interest Statement

The authors have declared no competing interest.

## References

Akaike, H. (1973). Information Theory and an Extension of the Maximum Likelihood Principle, pages 199–213. Springer New York, New York, NY.

Bi, Qifang et al. (2020). Epidemiology and transmission of COVID-19 in 391 cases and 1286 of their close contacts in Shenzhen, China: a retrospective cohort study. Lancet Infectious diseases, published online April 27, 2020.

Bliznashki, S. (2020). A Bayesian Logistic Growth Model for the Spread of COVID-19 in New York. Preprint from medRxiv, doi: 10.1101/2020.04.05.20054577

Boyce, W.E. & DiPrima, R.C. Elementary Differential Equations. 10th ed., Wiley (2012) Buizza, R. (2020). Probabilistic prediction of COVID-19 infections for China and Italy, using an ensemble of stochastically-perturbed logistic curves. arXiv preprint arXiv:2003.06418.

Cao, Y., Li, Q., Chen, J., Guo, X., Miao, C., Yang, H., Chen, Z., Li, C., & Li, L. (2020). Hospital Emergency Management Plan During the COVID-19 Epidemic. Academic Emergency Medicine 27(4), 309–311

Chen, Y., Li, Z., Zhang, Y. Y., Zhao, W. H., & Yu, Z. Y. (2020). Maternal health care management during the outbreak of coronavirus disease 2019 (COVID-19). Journal of Medical Virology, 92: 731–739.

Cheung, J. C. H., Ho, L. T., Cheng, J. V., Cham, E. Y. K., & Lam, K. N. (2020). Staff safety during emergency airway management for COVID-19 in Hong Kong. The Lancet Respiratory Medicine, 8(4).

Chowell, G. (2017). Fitting dynamic models to epidemic outbreaks with quantified uncertainty: a primer for parameter uncertainty, identifiability, and forecasts. Infectious Disease Modelling, 2(3), 379–398.

Coma, E., Mora, N., Prats-Uribe, A., Fina, F., Prieto-Alhambra, D., & Medina-Peralta, M. (2020). Excess cases of influenza suggest an earlier start to the coronavirus epidemic in Spain than official figures tell us: an analysis of primary care electronic medical records from over 6 million people from Catalonia. Preprint from medRxiv, doi: 10.1101/2020.04.09.20056259

Cribari-Neto, F., & Zeileis, A. (2009). Beta regression in R. Research Report Series, Department of Statistics and Mathematics 98, WU Vienna University of Economics and Business.

Dong, E., Du, H., & Gardner, L. (2020). An interactive web-based dashboard to track COVID-19 in real time. The Lancet Infectious Diseases, 20(5).

Ferrari, S. & Cribari-Neto, F. (2004). Beta Regression for Modelling Rates and Proportions. Journal of Applied Statistics, 31(7), 799–815.

Fox, J. & Monette, G. (1992). Generalized collinearity diagnostics. Journal of the American Statistical Association 87(417), 178–183.

Gange, S. J., Muñoz, A., Sáez, M., & Alonso, J. (1996). Use of the beta-binomial distribution to model the effect of policy changes on appropriateness of hospital stays. Journal of the Royal Statistical Society: Series C (Applied Statistics), 45(3), 371–382.

Grange, E. S., Neil, E. J., Stoffel, M., Singh, A. P., Tseng, E., Resco-Summers, K., Fellner, B. J., Lynch, J. B., Mathias, P. C., Mauritz-Miller, K., Sutton, P. R., & Leu, M. G. (2020). Responding to COVID-19: The UW Medicine Information Technology Services Experience. Applied clinical informatics, 11(2), 265–275.

Huang, R., Liu, M., & Ding, Y. (2020). Spatial-temporal distribution of COVID-19 in China and its prediction: A data-driven modeling analysis. The Journal of Infection in Developing Countries, 14(03), 246–253.

Hunger, M., Döring, A., & Holle, R. (2012). Longitudinal beta regression models for analyzing health-related quality of life scores over time. BMC medical research methodology, 12(1), 144.

Ivorra, B., Ferrández, M. R., Vela-Pérez, M., & Ramos, A. M. (2020). Mathematical modeling of the spread of the coronavirus disease 2019 (COVID-19) taking into account the undetected infections. The case of China. Communications in nonlinear science & numerical simulation, 88, 105303.

Liu, Y., Gayle, A. A., Wilder-Smith, A., & Rocklöv, J. (2020). The reproductive number of COVID-19 is higher compared to SARS coronavirus. Journal of travel medicine, 27(2).

Ministerio de Sanidad (2020). Evolución diaria de la pandemia de COVID-19 en España. Spanish Ministry of Health, Consumer Affairs and Social Welfare. Available at: https://covid19.isciii.es/ (xData retrieved on April 22nd, 2020)

Modi, C., Boehm, V., Ferraro, S., Stein, G., & Seljak, U. (2020). Total COVID-19 Mortality in Italy: Excess Mortality and Age Dependence through Time-Series Analysis. Preprint from medRxiv, doi: 10.1101/2020.04.15.20067074

Moraes, R. M., Rocha, A. V., & Machado, L. S. (2012). Intelligent assessment based on beta regression for realistic training on medical simulators. Knowledge-Based Systems, 32, 3–8.

Richards, F. J. (1959). A flexible growth function for empirical use. Journal of experimental Botany, 10(2), 290–301.

Schwarz, G. (1978). Estimating the dimension of a model. Ann. Statist., 6(2):461–464.

Villalobos-Arias, M. (2020). Estimation of population infected by COVID-19 using regression Generalized logistics and optimization heuristics. arXiv preprint arXiv:2004.01207.

Vollmer, S. & Bommer, C. (2020). Average detection rate of SARS-CoV-2 infections is estimated around nine percent. Preprint http://www.uni-goettingen.de/en/606540.html

Wu, K., Darcet, D., Wang, Q., & Sornette, D. (2020). Generalized logistic growth modeling of the COVID-19 outbreak in 29 provinces in China and in the rest of the world. arXiv preprint arXiv:2003.05681.

Yu, J., Ding, N., Chen, H., Liu, X. J., He, W. J., Dai, W. C., Zhou, Z. G., Lin, F., Pu, Z. H., Li, D. F., Xu, H. J., Wang, Y. L., Zhang, H. W., & Lei, Y. (2020). Infection Control against COVID-19 in Departments of Radiology. Academic radiology, 27(5), 614–617.

Zhang, Z. (2016). Parametric regression model for survival data: Weibull regression model as an example. Annals of translational medicine, 4(24).

